# Bridging Cultures to Defeat COVID-19: An Innovative Virtual Exchange Program in Global Medical Education

**DOI:** 10.1101/2024.09.30.24314434

**Authors:** Christina D. Campagna, Madison P. Searles, Joanna L. Suser, Ruwida M. K. Omar, Hanan Bugaigis, Khadiga Muftah Hilal Mansur, Abdelqader Imragaa, Nihar Ranjan Dash, Basema Saddik, Hani Shennib, Lawrence S. Chin, Seth W. Perry

## Abstract

**The problem and opportunity:** There is a critical and growing need to train globally focused, culturally fluent clinicians and scientists who can collaboratively defeat current and future public health threats across international boundaries. In parallel, academic conferences bring together thousands of diverse international healthcare professionals every year, yet their potential to provide the crucial professional development training necessary to advance internationalized medicine is often underutilized.

**The solution:** We developed and now first report an innovative healthcare education program that used an academic conference as the framework around which to build a structured, non-incidental virtual exchange (VE) for training globally and culturally proficient healthcare professionals. Herein we further describe the program’s design and content, successes and challenges, and lessons learned.

**Program Overview:** Using a smartphone based social-networking and conference management app with available translation capabilities, pre- and post-graduate trainees prepared and participated in poster presentations, seminars, and workshops to learn current research and best-practices in COVID-19 medicine, while engaging with their international peers in networking and professional-development exercises. The 2-week intensive program included daily synchronous interactive seminars on various topics in COVID-19 medicine, international team-based asynchronous activities such as preparing, presenting, and constructively critiquing research posters at virtual poster sessions, and expert-led wellness and cultural-competence workshops. Participants received initial training in the norms of intercultural communication, syllabus content and expectations, incentives, icebreaker activities, and program technology. They learned then-current COVID-19 medical research, therapies, and best practices, as well as professional "soft skills" including leadership, team building, scientific/clinical presentation, verbal/written communication skills, and intercultural competence. The program vastly expanded participants’ international professional networks to enhance their mentorship and career development opportunities.

**Conclusions:** Participants reported receiving substantial benefits from the program, with many reporting immediate translation of lessons learned toward improving healthcare education or practice in their home communities.

**TEASER:** Widespread innovative use of academic conferences as vehicles for structured non-incidental virtual exchange, professional development, and global medical education could improve healthcare education, capacity, and outcomes worldwide.

**KEY MESSAGES:**

- We developed and piloted a novel virtual exchange modality to connect international health science trainees and practitioners for unique collaborative training opportunities.
- Our "nested virtual exchange" concept employed an academic conference framework as the vehicle for providing structured cross-national didactics and professional development activities.
- This model aims to train a globally proficient next generation of clinicians and scientists who are optimally equipped to tackle current and future global health concerns.
- Our highly scalable, flexible, and efficient model can be adapted to any scientific or medical topic or focus, and is suitable for in-person, virtual, or hybrid approaches. It is especially suitable for student/trainee-led initiatives.
- Widespread adoption of this innovative training approach by universities, professional societies, and conference planners worldwide would equip many more healthcare providers and scientists with the knowledge and skills required to tackle public health challenges across international boundaries, thus improving global health outcomes.
- We hope that other universities, conference planners, and especially students and trainees will accept the baton to develop and launch similar programs to expand internationalized science and medicine worldwide.

## INTRODUCTION

The COVID-19 pandemic reshaped medical education worldwide, prompting an urgent shift to online learning and suspension of global health exchanges to safeguard students and communities. Although challenging^1^, these disruptions spurred educators to embrace innovative approaches for globalizing medical education. These innovations included rapid expansion of virtual learning opportunities, leveraging the intersection of globalization and technological advancements. While online learning has long been recognized as an essential delivery modality for global medical education^2^, its integration into academic global health programs has historically been limited^3^. The pandemic catalyzed demand for virtual learning experiences that could improve global health understanding, cultural competency, and foster peer connections. This ongoing shift signifies a pivotal moment wherein virtual learning is anticipated to play a key role in the future of global health education, thanks to its adaptability and ability to transcend geographical barriers.

Internationalized medical education is critical to enhancing trainees’ ability to practice medicine in today’s globalized world^4^. Curriculum internationalization involves intentionally integrating intercultural and global dimensions into post-secondary education to enhance quality and prepare trainees to contribute meaningfully to society^5^. By exposing students to diverse global populations, curriculum internationalization is pivotal for fostering cultural competence and professional development. This exposure helps students better understand social, cultural, and ethnic differences among the communities they will serve as healthcare professionals^4^. The COVID-19 pandemic underscored the urgent need to prepare a globally proficient generation of clinicians and scientists who can capably tackle current and future global health challenges. Educators have long advocated for advancing professional competencies, emphasizing the importance of communication across diverse cultural backgrounds^6,7^. An internationalized curriculum hones employability and prepares students with technical skills essential for navigating contemporary society including teamwork, negotiation, problem-solving, and communication^8^. Healthcare practitioners trained in cultural competence and humility can deliver high-quality care, improve patient outcomes, and mitigate health disparities among culturally diverse patient populations^9,10^.

This paper describes a novel academic conference that leveraged virtual exchange (VE) with an internationalized curriculum on scientific and medical approaches to defeat COVID-19. The *Bridging Cultures to Defeat COVID-19* (BCTDC19) virtual program offered collaborative academic-style lectures, professional development, and cultural exchange opportunities for trainees and faculty from the United States (US), Libya, and the United Arab Emirates (UAE). Given the demands of our globalized world coupled with the complexity of the COVID-19 pandemic, the need to foster dialogue and international collaboration to solve global health problems has never been greater.

A unique and innovative aspect of this VE learning experience was its implementation "within” an academic conference model. Academic conferences attract thousands of international participants yearly. During the COVID-19 pandemic, most conferences quickly went virtual. This created a tremendous untapped opportunity to exponentially grow VE’s reach by using academic conferences as vehicles around which to build meaningful online international learning experiences. This model simultaneously advances several global health missions, including curriculum internalization, providing critical scientific and professional development training to international medical trainees, and promoting cultural and global competence.

Few studies describe an experience like our BCTDC19 conference, a concise, two-week online peer-collaborative learning initiative. Existing studies have focused on longer-term programs like a 10-week peer-to-peer global health partnership^11^ and 5-week virtual global health case conferences^12^. These programs emphasized cross-cultural learning between students from different socio-economic backgrounds to enhance cultural competence and professional growth. Some studies have discussed the benefits of e-learning and experiential education in global health contexts^13,14^, with peer or collaborative learning components. The most comparable study involved a two-week reciprocal intercultural peer e-learning activity culminating in collaborative research with an international counterpart^6^.

A systematic review focusing on virtual global health in graduate medical education discussed the “crucial need for evidence about virtual global health education activities planning, implementation, and continuation… to guide global health educators and the creation of global health programming”^15^. Echoing this sentiment, our descriptive case study herein addresses this gap by detailing the goals, activities, structure, and practical considerations of implementing a virtual peer-collaborative learning and cultural exchange experience for scientific and medical trainees. It offers insights and lessons learned to guide future global health educators in designing similar initiatives in virtual medical education.

## PROGRAM DESCRIPTION

### Ethics Statement

The SUNY Upstate Medical University Institutional Review Board (IRB) determined that this project is exempt from IRB review.

### Conference Goals

BCTDC19 was a virtual health sciences education program that gathered American, Middle Eastern, and North African clinical and scientific trainees for collaborative scientific and medical education, and unique professional development and cultural exchange opportunities. Participants were undergraduate, graduate, and young adult (≥ 18 years old) pre- and post-degree health science trainees. International faculty and staff from various health science education programs provided the trainees with seminars/talks, instruction, mentorship, and networking opportunities.

### International Collaborations

Institutions with health sciences programs from Libya, the UAE, and the US were invited to participate. (Future programs not constrained by funding requirements would seek broader geographic diversity.) Employing a "hub and spokes" model, each country had a designated institution or non-governmental organization (NGO) overseeing regional recruitment of university partners, supervising/organizing faculty and staff, keynote speakers, and trainee participants: the National Council on US-Libya Relations (NCUSLR) for Libya, the Sharjah Research Technology and Innovation Park (SRTIP) for the UAE, and SUNY Upstate Medical University for the USA and overseeing all program operations. Having these trusted liaisons with deep knowledge of the local cultures, broad connections, and longstanding histories of respected and successful capacity building in the region was instrumental to overcoming the enormous logistical challenges and ensuring the success of BCTDC19. Trainees and faculty/staff from all regions and partner Institutions played crucial roles in coordinating and executing program activities, with diverse international trainees taking the lead wherever possible, under appropriate supervision. Program activities including talks, seminars, and other educational components were delivered by faculty and staff representing each participating country (**Figure S1**).

### Conference Structure

BCTDC19 occurred for two weeks in Fall 2021, and again in Spring 2022. The fully virtual conference provided educational sessions and activities to foster international and intercultural awareness, dialogue, and collaboration toward tackling global health problems. Daily live (virtual) keynote talks featured diverse international medical and public health experts. Journal clubs and social hours provided opportunity for group discussions and interactions, on themes that mirrored the daily keynote topic(s). BCTDC19 also offered professional development, cultural competence, wellness, bias and imposter syndrome, and medical ethics workshops. Cross-national trainee teams worked together on virtual journal club activities, preparing and presenting posters, and offering constructive feedback. Gamification activities enabled individuals and teams to earn participation points, with a leaderboard displayed on the virtual platform throughout the conference. Certificates, CME credits, and other non-monetary incentives were offered. Supplemental Figure 1 (**Figure S1**) shows the Spring 2022 conference flyer and agenda. Fall 2021 conference evaluations revealed areas of improvement and interest that were implemented for the Spring 2022 conference.

### Conference Certificate and Trainee Deliverables

Trainee participants wanting a conference completion certificate needed to engage in ≥ 10 hours on the conference platform, post five photos to the social wall, complete the pre- and post-conference surveys, and participate in ≥ 1 social hour and ≥ 1 academic activity during the two-week conference. To complete the academic activity, teams of 2-4 from different countries collaboratively created then presented posters on COVID-19 science or medicine, including peer-reviewed literature from all team members’ home country. Participants could also present their own prior research at the synchronous virtual poster session.

### Participatory Model of Trainee Engagement

A key element of BCTDC19 was having internationally diverse trainees collaboratively engaged as conference planners, facilitators, moderators, and near-peer mentors. Trainees from SUNY Upstate and international partner Institutions helped plan and implement the social and networking activities, gamification, and daily themes to promote participant engagement and cross-cultural connections and understanding. Trainees also served as moderators for keynote presentations, and facilitators for daily social hours and journal clubs. They engaged with conference attendees by posting on the social wall, and served as teammates for the peer-poster deliverable. Trainee facilitator and participant feedback on areas for program improvement and interest guided the addition of new/improved content for the Spring 2022 session.

### Trainee Recruitment and Participation

Trainee participants were pre- and post-graduate health sciences trainees from the US, Libya, and the UAE. Participation was geographically restricted by funding requirements, but otherwise any interested health or science trainee ≥ 18 years old could participate. Most trainee participants came from medicine, pharmacy/pharmacology, and public health (**Table 1**). Conference recruitment occurred through outreach campaigns led by faculty/staff program organizers in the target countries.

**Table 1:** Participant Data for Bridging Cultures to Defeat COVID-19. **Conference Participant Data for Fall 2021 and Spring 2022**

| **Table 1: Conference Participant Data for Fall 2021 and Spring 2022** |  |  |  |  |
| --- | --- | --- | --- | --- |
| **Characteristic** | **Fall 2021** |  | **Spring 2022** |  |
|  | **n** | **%** | **n** | **%** |
| **New Trainee Responses (Fall 2021 and Spring 2022) n=247 Fall, n=191 Spring** |  |  |  |  |
| **Age mean (SD)** | 26.56 (5.90) |  | 26.99 (7.88) |  |
| **Gender** | **n** | **%** | **n** | **%** |
| Male | 82 | 33.2 | 53 | 27.7 |
| Female | 162 | 65.6 | 135 | 70.7 |
| Non-Binary | 2 | 0.8 | 1 | 0.5 |
| Prefer not to answer | 1 | 0.4 | 2 | 1.0 |
| **University Country** |  |  |  |  |
| Libya | 106 | 43.6 | 87 | 45.5 |
| United Arab Emirates | 25 | 10.3 | 39 | 20.4 |
| United States | 96 | 39.5 | 58 | 30.4 |
| Other | 16 | 6.6 | 7 | 3.7 |
| **Current Academic Institution** |  |  |  |  |
| University of Benghazi | 43 | 17.4 | 56 | 29.3 |
| National Arab American Medical Association (NAAMA) | 1 | 0.4 | 0 | 0.0 |
| National Council on US-Libya Relations (NCUSLR) | 1 | 0.4 | – | – |
| Sharjah Research Technology and Innovation Park (SRTIP) | – | – | 1 | 2.9 |
| SUNY Upstate Medical University | 54 | 21.9 | 8 | 22.9 |
| University of Sharjah | 24 | 9.7 | 0 | 0.0 |
| University of Tripoli | 44 | 17.8 | 7 | 20.0 |
| University of Zawia | 19 | 7.7 | 1 | 2.9 |
| Other | 61 | 24.7 | 3 | 8.6 |
| **Institution Funding** |  |  |  |  |
| Private | 40 | 16.2 | 42 | 22.0 |
| Public | 207 | 83.8 | 146 | 76.4 |
| **English is Primary Language at Institution** |  |  |  |  |
| Yes | 229 | 92.7 | 181 | 94.8 |
| No | 18 | 7.3 | 10 | 5.2 |
| **Highest Degree Earned** |  |  |  |  |
| Bachelors | 123 | 49.8 | 94 | 49.2 |
| Masters | 34 | 13.8 | 19 | 9.9 |
| MD/MBBS | 31 | 12.6 | 18 | 9.4 |
| PhD | 3 | 1.2 | 5 | 2.6 |
| DrPh | 0 | 0.0 | 1 | 0.5 |
| Other | 56 | 22.7 | 54 | 28.3 |
| **Level (Year) of Training in Current Program** |  |  |  |  |
| Year 1 | 75 | 18.3 | 51 | 28.2 |
| Year 2 | 47 | 19.0 | 20 | 11.0 |
| Year 3 | 21 | 8.5 | 26 | 14.4 |
| Year 4 | 52 | 21.1 | 50 | 27.6 |
| Year 5 | 34 | 13.8 | 24 | 13.3 |
| Year 6 | 18 | 7.3 | 10 | 5.5 |
| **Top Six Current Education Programs** |  |  |  |  |
| Dentistry | 10 | 5.1 | – | – |
| Medicine | 83 | 42.3 | 111 | 73.5 |
| Neurology | 11 | 5.6 | – | – |
| Pharmacy/Pharmacology | 19 | 9.7 | 13 | 8.6 |
| Public Health | 20 | 10.2 | 5 | 3.3 |
| Radiation Therapy | 6 | 3.1 | – | – |
| **Alumni Trainee Responses (Spring 2022 only)**<br>**n=35** |  |  |  |  |
| **Age mean (SD)** | – | – | 28.03 (4.37) |  |
| **Gender** |  |  |  |  |
| Male | – | – | 8 | 22.9 |
| Female | – | – | 27 | 77.1 |
| Non-Binary | – | – | 0 | 0.0 |
| Prefer not to answer | – | – | 0 | 0.0 |
| **University Country** |  |  |  |  |
| Libya | – | – | 24 | 68.6 |
| United Arab Emirates | – | – | 1 | 2.9 |
| United States | – | – | 8 | 22.9 |
| Other | – | – | 2 | 5.7 |
| **Current City** |  |  |  |  |
| Benghazi | – | – | 15 | 42.9 |
| Syracuse | – | – | 8 | 22.9 |
| Tripoli | – | – | 8 | 22.9 |
| **Current Academic Institution** |  |  |  |  |
| University of Benghazi | – | – | 14 | 40.0 |
| National Arab American Medical Association<br>(NAAMA) | – | – | 1 | 2.9 |
| National Council on US-Libya Relations<br>(NCUSLR) | – | – | – | – |
| Sharjah Research Technology and Innovation<br>Park (SRTIP) | – | – | 1 | 2.9 |
| SUNY Upstate Medical University | – | – | 11 | 22.9 |
| University of Sharjah | – | – | 0 | 0.0 |
| University of Tripoli | – | – | 7 | 20.0 |
| University of Zawia | – | – | 1 | 2.9 |
| Other | – | – | 3 | 8.6 |
| **Institution Funding** |  |  |  |  |
| Private | – | – | 0 | 0.0 |
| Public | – | – | 35 | 100.0 |
| NGO institution | – | – | 0 | 0.0 |
| **English is Primary Language at Institution** |  |  |  |  |
| Yes | – | – | 33 | 94.3 |
| No | – | – | 2 | 5.7 |
| **Highest Degree Earned** |  |  |  |  |
| Bachelors | – | – | 15 | 42.9 |
| Masters | – | – | 9 | 25.7 |
| MD/MBBS | – | – | 2 | 5.7 |
| PhD | – | – | 1 | 2.9 |
| DrPh | – | – | 1 | 2.9 |
| Other | – | – | 7 | 20.0 |
| **Level (Year) of Training in Current Program** |  |  |  |  |
| Year 1 | – | – | 5 | 15.6 |
| Year 2 | – | – | 7 | 21.9 |
| Year 3 | – | – | 1 | 3.1 |
| Year 4 | – | – | 12 | 37.5 |
| Year 5 | – | – | 7 | 21.9 |
| Year 6 | – | – | 0 | 0.0 |
| **Faculty/Staff Responses (Fall 2021 and Spring 2022) n=163 Fall 2021, n=157 Spring 2022** |  |  |  |  |
| **Gender** |  |  |  |  |
| Male | 56 | 34.4 | 46 | 29.3 |
| Female | 106 | 65.0 | 109 | 69.4 |
| Non-Binary | 1 | 0.6 | 0 | 0.0 |
| Prefer not to answer | 0 | 0.0 | 2 | 1.3 |
| **Role at Conference** |  |  |  |  |
| Faculty/Staff Participant | 110 | 81.5 | 126 | 80.3 |
| Faculty/Staff Presenter | 19 | 14.1 | 31 | 19.7 |
| Both Faculty/Staff Participant and Presenter | 6 | 4.4 | – | – |
| **Country of Employment** |  |  |  |  |
| Libya | 41 | 25.9 | 27 | 17.2 |
| United Arab Emirates | 8 | 5.1 | 27 | 17.2 |
| United States | 99 | 62.7 | 89 | 56.7 |
| Other | 10 | 6.3 | 14 | 8.9 |
| **Top four Current Cities** |  |  |  |  |
| Ajman | – | – | 13 | 8.4 |
| Benghazi | 19 | 12.0 | 14 | 9.1 |
| Syracuse | 66 | 41.8 | 55 | 35.7 |
| Tripoli | 10 | 6.3 | 12 | 7.8 |
| Zawia | 9 | 5.7 | – | – |
| **Current Academic Institution** |  |  |  |  |
| University of Benghazi | 22 | 13.6 | 14 | 8.9 |
| National Arab American Medical Association (NAAMA) | 0 | 0.0 | 0 | 0.0 |
| National Council on US-Libya Relations (NCUSLR) | 1 | 0.6 | – | – |
| Sharjah Research Technology and Innovation Park (SRTIP) | – | – | 0 | 0.0 |
| SUNY Upstate Medical University | 63 | 38.9 | 63 | 40.1 |
| University of Sharjah | 2 | 1.2 | 7 | 4.5 |
| University of Tripoli | 11 | 6.8 | 12 | 7.6 |
| University of Zawia | 10 | 6.2 | 0 | 0.0 |
| Other | 53 | 32.7 | 61 | 38.9 |
| **Institution funding** |  |  |  |  |
| Private | 14 | 8.9 | 21 | 13.4 |
| Public | 129 | 82.2 | 124 | 79.0 |
| NGO institution | 1 | 0.6 | 2 | 1.3 |
| Other | 13 | 8.3 | 10 | 6.4 |
| **English is primary language at institution** |  |  |  |  |
| Yes | 154 | 94.5 | 146 | 93.0 |
| No | 9 | 5.5 | 11 | 7.0 |
| **Degree/Credentials** |  |  |  |  |
| Bachelors | 23 | 14.1 | 16 | 10.2 |
| Masters | 39 | 23.9 | 53 | 33.8 |
| MD/MBBS | 48 | 29.5 | 36 | 22.9 |
| PhD | 30 | 18.4 | 40 | 25.5 |
| MD/MBBS, PhD | 5 | 3.0 | 2 | 1.2 |
| Other | 18 | 11.0 | 10 | 6.4 |
| **Title at Current Institution** |  |  |  |  |
| C-Level Executive | 9 | 6.9 | — | — |
| Dean | 5 | 3.8 | — | — |
| Professor | 63 | 48.5 | 55 | 41.7 |
| Nurse | — | — | 8 | 6.1 |
| Physician | — | — | 5 | 3.8 |
| Program Director | — | — | 12 | 9.1 |

### Virtual Platform

The event hosting platform Socio (subsequently rebranded as Webex Events)^16^ connected learners and created an interactive event that fostered continuous attendee engagement. Socio was carefully chosen for its user-friendly interface and comprehensive event management capabilities, allowing seamless virtual event hosting and connection among attendees worldwide. Customizable event management features included a branded registration page, and schedulable email and push notifications for pre/mid-event reminders. To enhance the attendee experience, Socio offered customizable video rooms (used for the virtual journal clubs, social hours, and poster sessions), gamification tools, social wall, and chat features, and enabled fully embedded streaming and recording of all (virtual) live presentations (including Zoom integration). Engagement was encouraged through gamification, live Q&A sessions, and social wall prompts to stimulate participant conversation. Socio enabled collating attendee data including session attendance, connections made, and engagement statistics. BCTDC19 was accessible through web browsers or Socio’s dedicated smartphone application, ensuring seamless and equitable access to all content regardless of the chosen device. Together these technically advanced yet user-friendly features maximized program capabilities, engagement, and satisfaction.

### Registration

Registration occurred on the Socio platform. Participants chose a ticket type dependent on their participation status: Trainee or Faculty/Staff. There were no registration, tuition, or participation fees. Demographics were collected for each group and are summarized in **Table 1**.

## PROGRAM INSIGHTS AND LESSONS LEARNED

BCTDC19 was a fully virtual conference that gathered US and MENA health sciences trainees for collaborative scientific and medical education, professional development, and cultural exchange opportunities. This unique voluntary experience provided novel virtual learning experiences to clinical and non-clinical trainees from diverse fields, countries, and backgrounds, and fostered global connections despite pandemic-imposed social isolation. BCTDC19 focused on best practices and culturally relevant ways to reduce morbidity and mortality from COVID-19, but a similarly structured "nested virtual exchange" approach could be applied to virtual, in-person, or hybrid conferences within any field or topic.

Global health education promotes collaborative, multidisciplinary training practices to advance health equity for health concerns that are not “bound to a single geography or culture”^17^. VE represents a key growth area in global health education, given the critical need to grant low- and middle-income country (L&MIC) partners a seat at the table while providing equitable education opportunities for their learners^17^. [Note: herein we use "low- and middle-income country/ies" (L&MIC/L&MICs) to refer to, in aggregate, the World Bank’s classifications of low-income country (LIC), lower-middle income country (LMIC), and upper-middle income country (UMIC)^18,19^.] We must train health sciences students, both locally and globally, to control and treat current and emerging global health threats not endemic to their home region^6,20^.

### Conference Accessibility

Global health education currently strives to evolve from existing colonial models to ones of health equity. This evolution ensures that students, faculty, and trainees in L&MICs have equal programmatic input and access to educational resources compared to their high-income country (HIC) counterparts^21^. BCTDC19 promoted these ideals by actively involving faculty/staff and trainees from partner MENA schools in the post-award conference planning, prioritizing their needs and input. Further, US and MENA faculty experts and trainees co-participated in the keynote sessions, social hours, and journal clubs. This interactive engagement between faculty, staff, trainees, and stakeholders from each host country ensured equitable, multilateral collaboration. Furthermore, this approach helped to build trust, foster inclusivity, recognize and respect expertise, and build capacity with global partners.

Traditional global health programs often face access and cost barriers to trainee participation^22^. HIC students with sufficient means participate in global health programs more often than their peer counterparts in L&MICs^22^ Emerging best practices in global health education promote equality in partnerships where L&MIC trainees can train at HIC partner institutions^23^ The BCTDC19 conference had no registration, tuition, travel, or lodging fee barriers to participation.

Virtual learning also increases flexibility in work-life balance^24^. The BCTDC19 conference promoted such flexibility and minimized technology access and connectivity barriers by making all content accessible via both desktop browser and smartphone apps (iOS, Android). BCTDC19 content was also provided in multiple formats, such as synchronous (real-time) and asynchronous (recorded) keynote presentations, interactive live workshops, and live video chat rooms for social hours and journal clubs. Live activities were available at various times to accommodate different participant time zones and schedules. All presentations were recorded and uploaded to the platform, to accommodate those unable to (virtually) attend in real-time. With these features we strove to minimize participation barriers and maximize support for as many differing circumstances as possible, and provide equitable content access and opportunities for all HIC and L&MIC participants. BCTDC19 highlights the benefits and possibilities of making global health programming free and fully accessible to learners worldwide, and promoting global educational equity by limiting structural, social, and economic participation barriers.

### The Topic of COVID-19

The Socio online platform provided a space for discussing the globally impactful topic of COVID-19. Academic conferences on this subject help medical and health sciences trainees stay informed about the latest research, treatments, and guidelines in the rapidly evolving field. BCTDC19 featured diverse experts offering interdisciplinary perspectives on the pandemic’s medical, public health, and social dimensions. Keynote lectures delivered crucial emerging and evolving information on the virus, transmission, and treatment, thus helping trainees provide effective care to their home communities. The pandemic accelerated medical research, leading to treatment and healthcare delivery innovations. Learning about COVID-19 enabled trainees to stay updated on these advancements, and underscored the importance of public health measures and preparedness to protect communities. COVID-19 education was and remains essential for medical trainees to be well-prepared, informed, and adaptable global healthcare professionals.

### Educating the Global Healthcare Workforce Via Virtual Exchange

Motivations for promoting VE include enhancing educational capacity, fostering work-life flexibility, and ensuring cost-effectiveness of educational resources^24^ In response to resource limitations in L&MICs, virtual learning has been utilized to train the healthcare workforce due to faculty shortages, and to supplement existing formal teaching modalities^25,26^ These tools are especially important for capacity building in L&MICs, because they provide clinicians and researchers with real-time access to experts and information on treating difficult cases, managing morbidity and mortality, and adapting to the changing needs of their local community^26^. E-learning is advocated to overcome resource barriers in L&MICs with decentralized healthcare systems and limited training opportunities. BCTDC19 enhanced the Libyan healthcare system’s capacity by providing a cost-effective means of connecting trainees and faculty/staff with educational, professional development, and networking opportunities that might otherwise have been unavailable due to financial and/or geographic barriers. BCTDC19’s timely focus on COVID-19 education proved an excellent vehicle for meaningful discussion and collaboration among participants.

### Cultural Competency and Professional Development

The need to train future clinicians and scientists as culturally competent healthcare workers is increasingly evident ^9,10^. BCTDC19 improved international competencies through keynote presentations that shared medical research from diverse countries and universities. Social hours and journal clubs allowed participants to discuss topics such as how the pandemic has impacted their academic and home lives. Expert-led workshops focusing on imposter syndrome, biases, and wellness helped educate and empower students regarding common barriers faced in medicine. The Socio platform also provided key networking features such as direct and group messaging and video chats that allowed all participants to interact freely during and after BCTDC19, thus promoting global connection and collaboration.

The team-based poster and journal club preparations and presentations provided a structure for teamwork, negotiation, mediation, problem solving, and communication. Further, by requiring that the teams incorporate research from all team members’ home countries, these activities encouraged diverse perspectives and shared appreciation of international expertise. Preparing and delivering posters, journal clubs, talks, and papers are integral to academic training and life for most clinical and non-clinical trainees and professionals alike, and teach students how to understand and interpret medical and scientific topics, and effectively explain them to colleagues and peers. Despite the ubiquity of these activities within academia, many of our L&MIC trainee participants had no prior experience with (e.g.) poster or journal club preparations or presentations. While the same is likely true of many HIC health science students – depending on their age, program, and field – these anecdotes highlight probable disparities between the typical health science education experience in HICs versus L&MICs. Our program helped bridge that gap and advance equity by providing training experiences previously unavailable to many participants.

## CHALLENGES

Balancing the need for real-time interaction and accommodating participants’ varied schedules and time zones was a significant challenge for BCTDC19. To address this, all keynote presentations were delivered virtually in "real time" and recorded, so participants could either watch them "live" or review them asynchronously at their convenience. This approach helped to facilitate scholarly exchange and dialogue despite the scheduling difficulties.

Trainees could earn certificates by participating in a journal club and collaboratively creating and presenting a research poster with a trainee(s) from another country. However, limited certificate participation by HIC students reduced opportunities for international pairings, leading to some trainees being paired with students from different institutions within their home country. This observation suggested that HIC and L&MIC trainees may have had different goals and motivations for program participation and earning certificates, and/or simply different scheduling challenges.

Internet (Wi-Fi) and/or cellular connectivity or bandwidth issues sometimes posed challenges for speakers or participants (**Figure 1**). To mitigate these, we recorded presentations and made chat features available to all participants. The Socio iOS and Android apps were useful for many participants, allowing smartphone-based engagement when computer or Wi-Fi access was limited. Despite these challenges, participant feedback was overwhelmingly positive (**Figure 2**).

**Figure 1:**
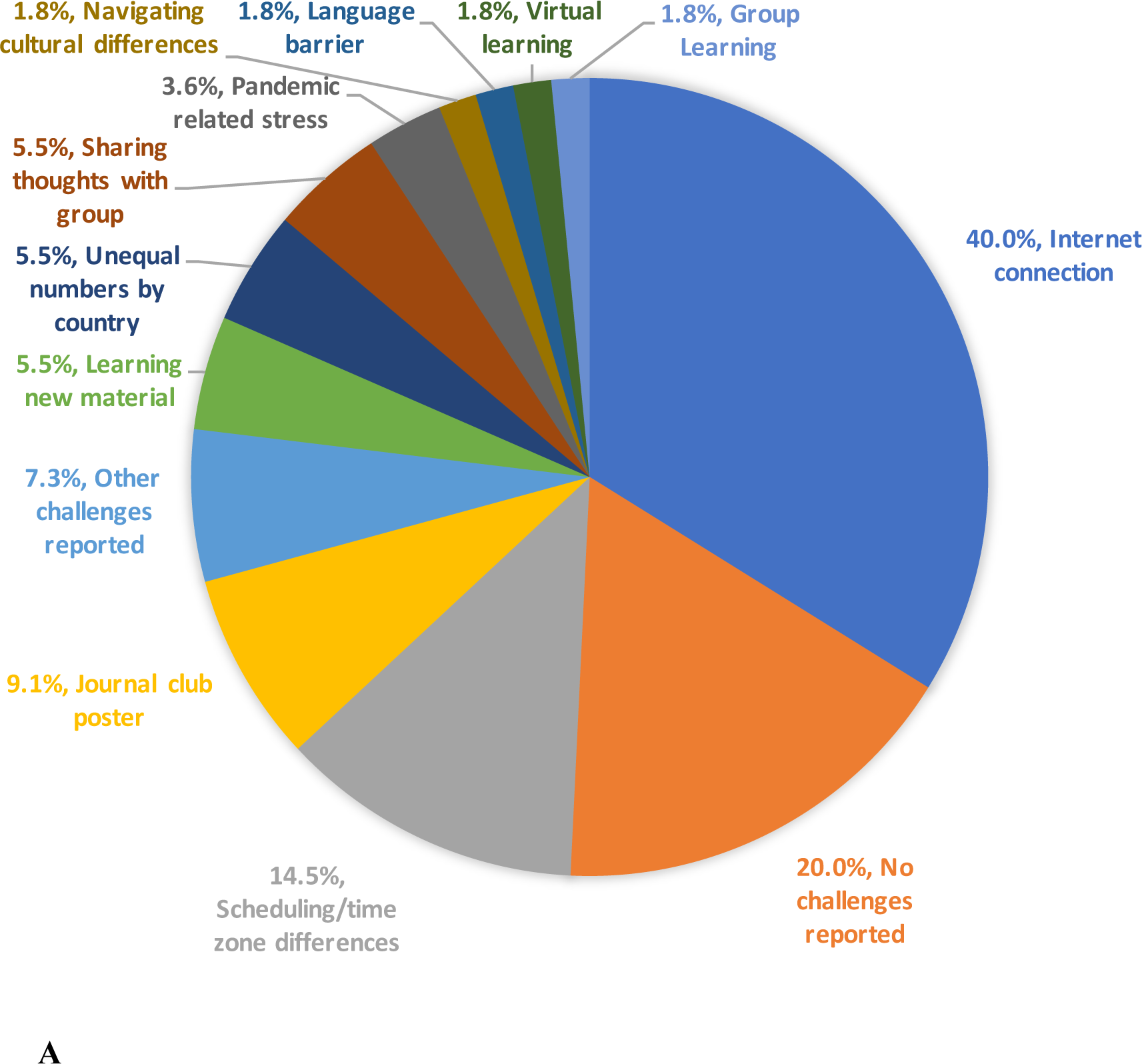
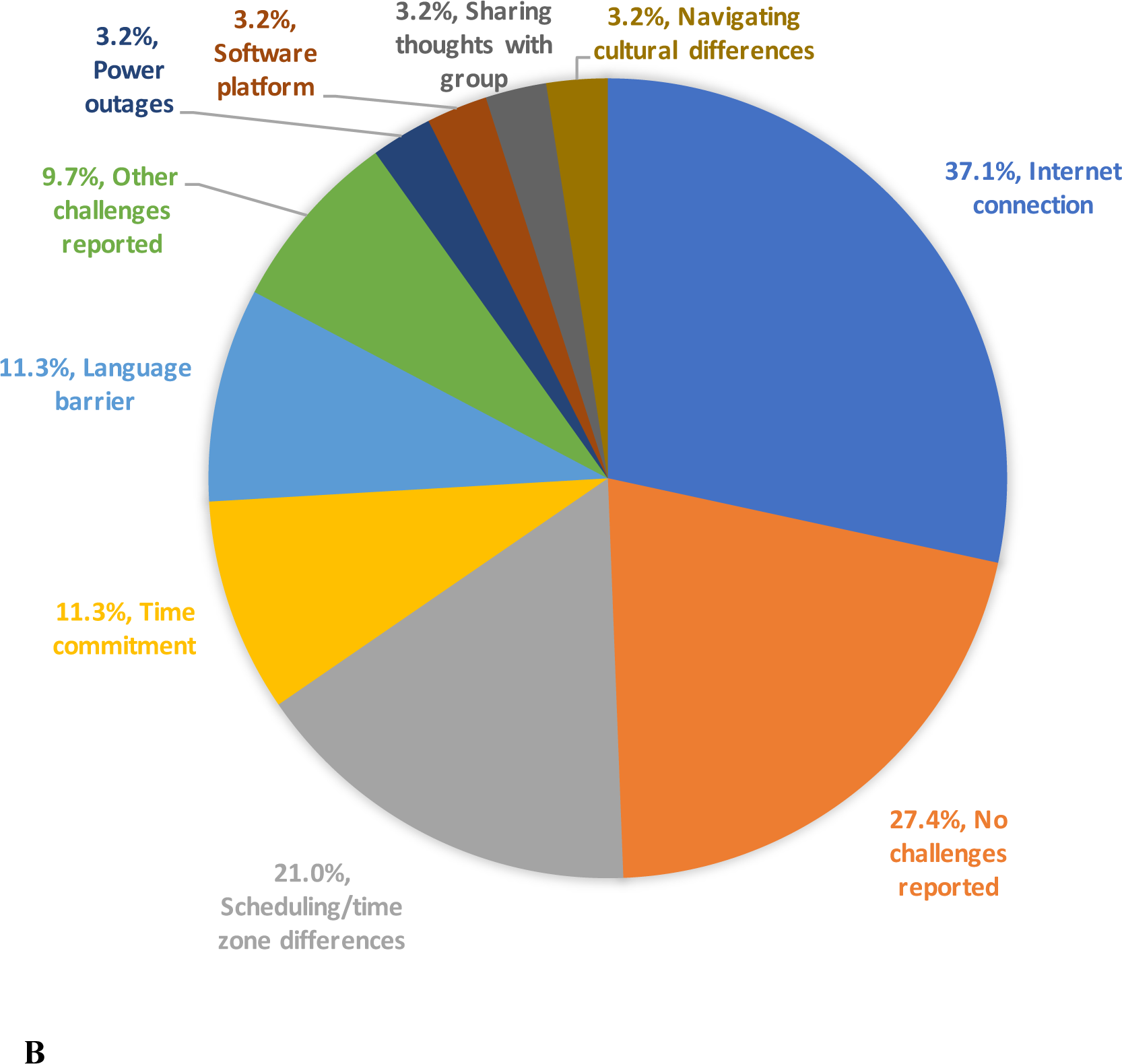
**A)**BCTDC19 Session 1 (Fall 2021) participants’ narrative responses to the post-program survey prompt "please comment on some of the challenges you experienced during your participation in the conference" were synopsized, aggregated categorically, and presented as the percentage of respondents citing the listed concern. Some respondents cited more than one concern, therefore the total percentage exceeds 100%. **B)** Same, for BCTDC19 Session 2 (Spring 2022).

**Figure 2:**
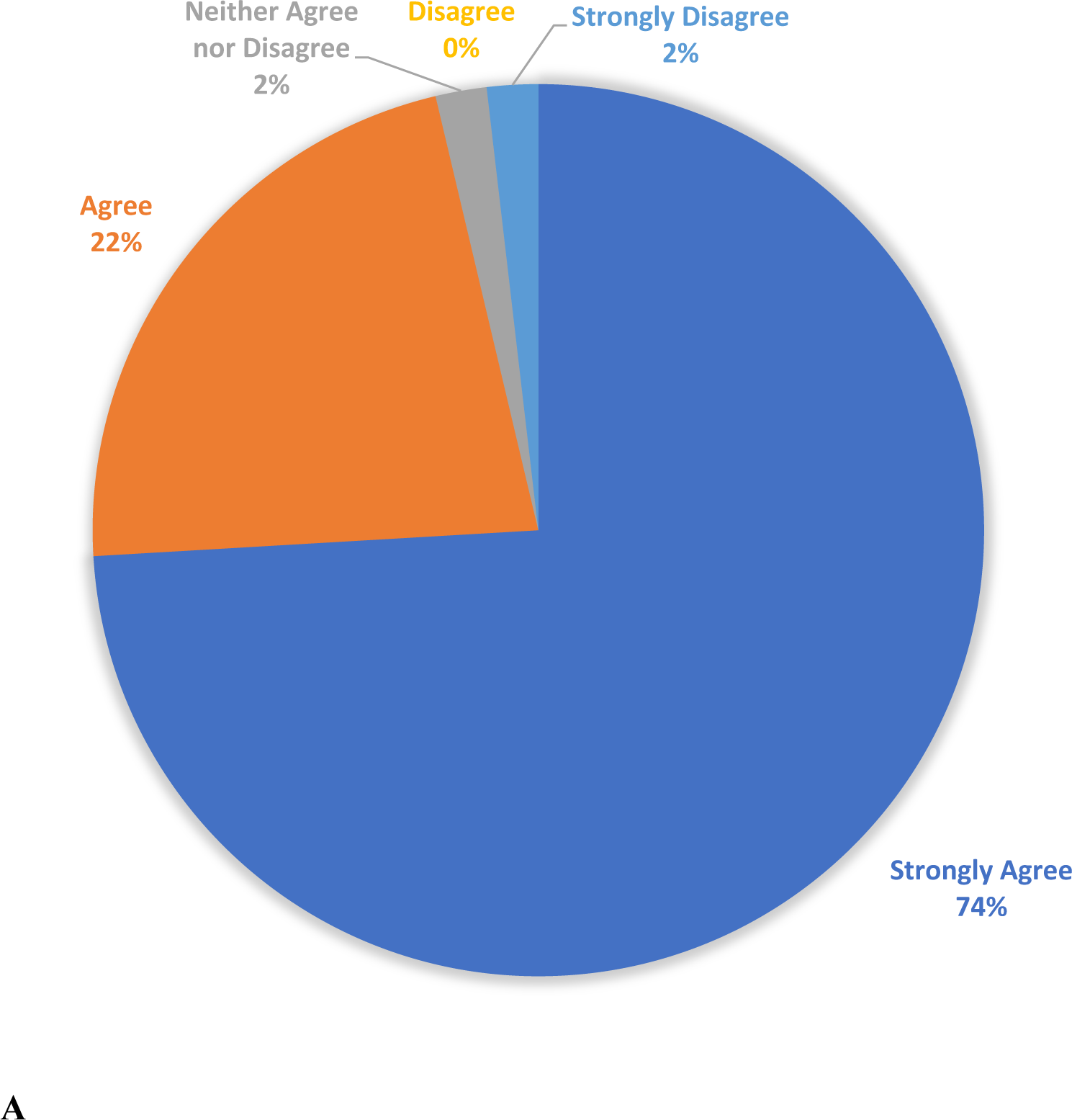
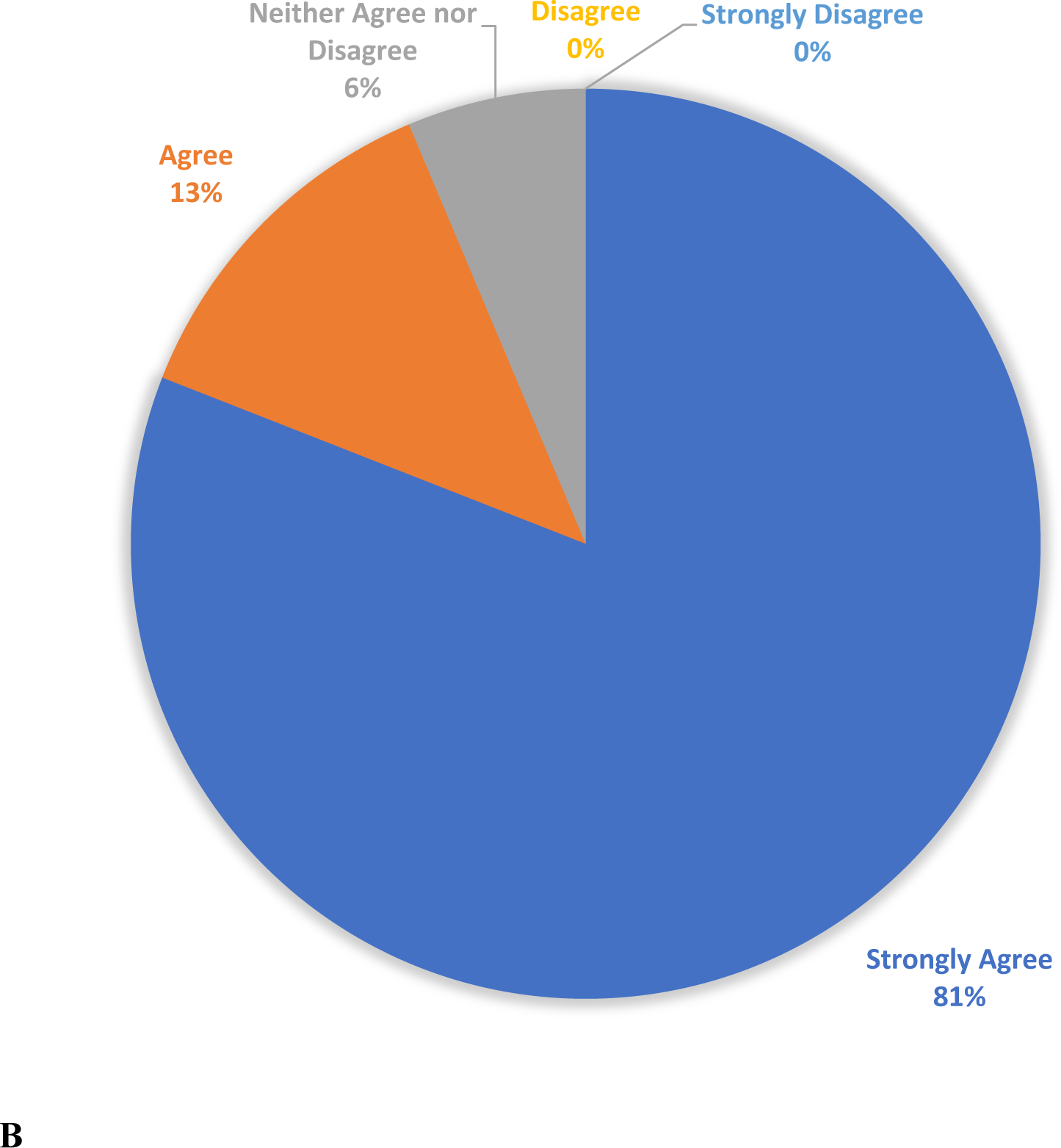
**A)** BCTDC19 Session 1 (Fall 2021) participant responses to the post-program survey prompt "I would recommend Bridging Cultures to Defeat COVID-19 to other students." Percentage of respondents selecting each of the five multiple choice answers are shown. **B)** Same, for BCTDC19 Session 2 (Spring 2022).

## CONCLUSIONS

The global health field has been developed, strengthened, and disseminated by the demands of our globalized world. The need to establish an internationalized curriculum that supports student learning in cultural competence, professional development, and interdisciplinary approaches to medicine has never been greater. Our experience building and administering BCTDC19 demonstrates that VE programs featuring internationalized curricula are valuable tools for training healthcare learners to evaluate, synthesize, and apply knowledge to prepare them for the real-world challenges of today and tomorrow.

Virtual learning plays crucial roles in disseminating strategic information on clinical care practices and expanding training globally. This approach addresses challenges in resource availability and contributes to overcoming limitations in educational infrastructure for trainees and faculty in HIC and L&MICs alike. BCTDC19 was a seminal virtual health sciences learning experience that featured structured and non-incidental VE within an academic conference, and designed collaborative learning experiences among international peers from HICs and L&MICs. The BCTDC19 structure is transferable to other academic institutions wishing to employ academic conferences as vehicles for hosting globally collaborative trainee learning experiences.

## Acknowledgements

The authors and SUNY Upstate Medical University profoundly thank the National Council on US Libya Relations, the University of Benghazi, the University of Tripoli, Sharjah Research Technology and Innovation Park, the University of Sharjah, and Zawia University, along with their participating faculty, staff, and students, for their invaluable assistance with local program recruitment, organization, implementation, and supervision, as well as their steadfast and unwavering Institutional support of program activities. We further thank the National Arab American Medical Association (NAAMA) and Mr. Ethan Chorin for their recruiting assistance and enthusiasm for this project. We also thank all student, faculty, and staff organizers, facilitators, presenters, mentors, and their Institutions from the US, Libya, and the UAE; and all conference participants. Their collective enthusiasm and assistance with participant and speaker recruitment, keynote presentations and facilitation, advertising, and similar regional conference logistics were vital to ensuring this program’s success. Last, but by no means least, we wish to express our sincere gratitude to the Stevens Initiative and their staff for supporting and believing in this project, and for all the incredible work they do each and every day to advance virtual exchange and improve lives around the world. The impact of their work in this area is immeasurable. Some text of this paper is excerpted with or without modifications from our "Bridging Cultures to Defeat COVID-19" grant application (unpublished work, copyright Seth W. Perry, 2020), with permission from the author.

## Author contributions

Conceptualization, program design: SWP, LSC, CDC, MPS; funding acquisition: SWP; recruitment, program administration/implementation/refinement, supervision, resources: all authors; data acquisition (registration and survey data), data curation, formal analysis, visualization: CDC, SWP, MPS, JLS; writing – original draft: CDC, SWP; writing – review & editing: all authors.

## Data Availability

All described data are included in the article and/or supplementary materials.

