## Supplemental Figure S1 for "Bridging Cultures to Defeat COVID-19: An Innovative Virtual Exchange Program in Global Medical Education"

#### February 28th - March 13th, 2022

##### Program Overview:

SUNY Upstate Medical University's Bridging Cultures to Defeat COVID-19 is a virtual health sciences education program that brings together American, Middle Eastern, and North African clinical and scientific trainees for collaborative scientific and medical education, and for unique professional development and cultural exchange opportunities.

##### Participants:

Undergraduate, Graduate, and Young Adult (> 18 years of age) pre- and post-degree health science trainees. Graduate and Medical faculty/staff are also encouraged to register and participate to provide mentorship and networking opportunities for the trainees. CME credit and professional development certificates will be available.

Click Here To Register

**How to Register:** [redacted]

**Questions:** [redacted]

##### Activities:

The conference will offer traditional academic and scientific activities intended to foster international and intercultural awareness, dialogue, and collaboration toward tackling global health problems. Trainee participants attend, and some may deliver, real-time or pre-recorded seminars and poster sessions on various topics of COVID-19 science and medicine.

##### Topics Covered Include:

- Epidemiology and Public Health
- Testing and Diagnostics
- Emergency and Critical Care
- Pediatric Disorders and Complications
- Neurological Symptoms and Complications
- Psychiatric and Mental Health Complications and Considerations
- Vaccine and Non-vaccine Therapeutic Strategies
- The New Normal: Preparing Hospital Operations to Safeguard Lives and Livelihoods

Stevens Initiative | aspen institute | UPSTATE MEDICAL UNIVERSITY | NCUSLR National Council on U.S. Libya Relations

Bridging Cultures to Defeat COVID-19 is supported by the Stevens Initiative, which is sponsored by the U.S. Department of State, with funding provided by the U.S. Government, and is administered by the Aspen Institute. The Stevens Initiative is also supported by the Bezos Family Foundation and the governments of Morocco and the United Arab Emirates. Additional support for Bridging Cultures to Defeat COVID-19 is provided by the State University of New York (SUNY) Upstate Medical University, the National Council on US Libya Relations (NCUSLR), Sharjah Research Technology and Innovation Park (SRTIP), the University of Benghazi, the University of Tripoli, Zawia University, the University of Sharjah, the National Arab American Medical Association (NAAMA), and the Nappi Institute of SUNY Upstate Medical University.

### Bridging Cultures to Defeat COVID-19 Virtual Exchange Conference

#### Team Building & Gamification:

International teams will engage and work together in virtual journal club activities, preparing posters or presentations, and offering constructive feedback. Individuals and teams will have the opportunity to earn points for participation through gamification activities, with a leaderboard displayed throughout the event on the virtual platform.

#### Professional Development:

Guided cultural competence building and awareness, implicit bias, and imposter syndrome exercises and workshops will be offered. Training will be led and designated by [redacted]

#### Incentives:

- Network and learn with experienced professionals in your field.
- Trainees who meet the required program criteria may request digital completion certificates/badges.

#### Learning Objectives:

- Enhance professional development through networking and reflecting with participants from other countries.
- Use effective cultural communication strategies when interacting with others.
- Demonstrate an understanding of the healthcare needs and disparities of diverse populations.
- Discuss how cultural beliefs shape interpretations and experience of disease(s) and treatment(s) across the world.
- Understand and utilize strategies and resources to instill cultural competence as a life-long learning process.
- Demonstrate an understanding of the roles and responsibilities of healthcare, public health, and research professionals as part of an interprofessional team.
- Prepare learners to synthesize, evaluate, and apply knowledge to tackle current and future global health concerns collaboratively and equitably.
- Use technology, information resources, and diverse ways of knowing to solve problems through virtual exchange.

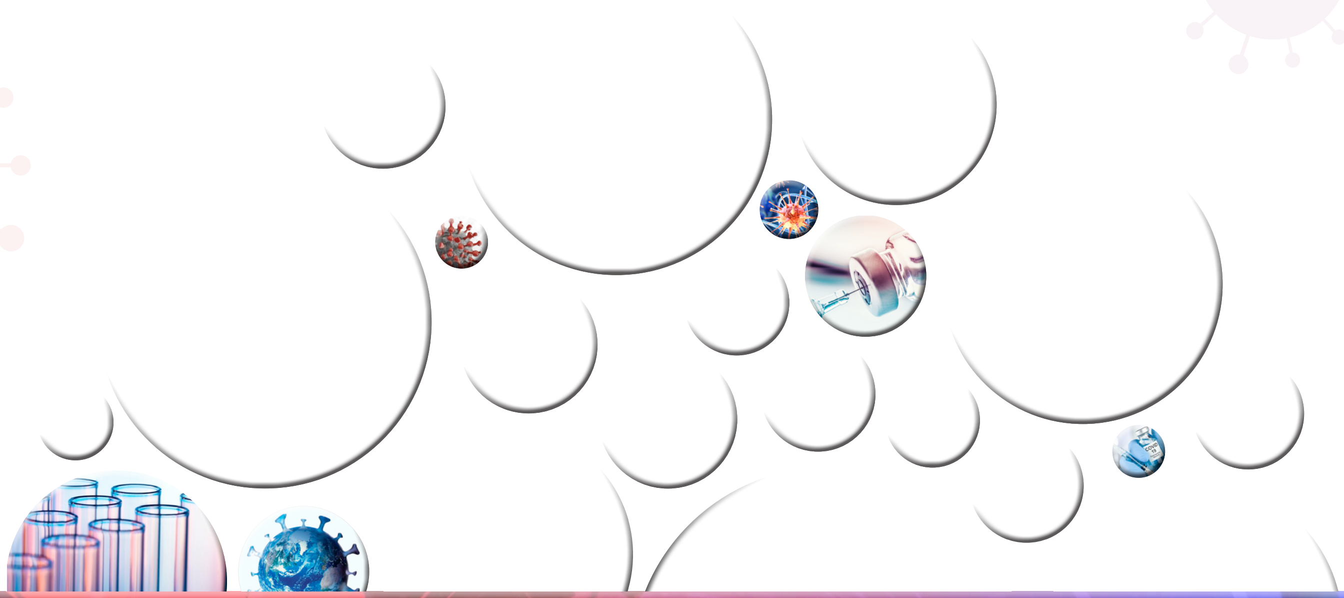

Any and all program details and information conveyed herein is not guaranteed and is subject to change.
This is not a comprehensive list of all program details or expectations. Program details or expectations may be added, removed, or altered at any time, at the sole discretion of the program organizers and sponsors.

**Bridging Cultures to Defeat COVID-19 Virtual Conference**
**Spring Agenda, February 28 – March 13, 2022**

#### Monday, February 28th

| | |
|---|---|
| | **Daily Theme: Introductions** |
| 7:00 – 7:15 AM EST | **Opening Remarks**<br>[redacted]<br>*Upstate Medical University* |
| 7:15 – 8:45 AM EST | **Keynote Panel: Epidemiology and Public Health**<br>[redacted] |
| 9:00 – 10:00 AM EST | **Keynote: Pediatrics and the COVID-19 Pandemic**<br>[redacted] |
| 12:00 – 1:00 PM EST | **Social Hour: Conference Welcome** |

#### Tuesday, March 1st

| | |
|---|---|
| | **Daily Theme: Commute to School** |
| 7:00 – 8:30 AM EST | **Keynote Panel: Neurological Complications of COVID-19**<br>[redacted] |
| 11:00 – 11:45 PM EST | **Journal Club: COVID-19: from Acute to Chronic Disease? Potential Long-Term Health Consequences** |
| 12:00 – 1:30 PM EST | **Workshop: Wellness**<br>[redacted] |

#### Wednesday, March 2nd

| | |
|---|---|
| | **Daily Theme: Academic Papers** |
| 7:00 – 8:30 AM EST | **Keynote Panel: Pediatric Disorders and Complications**<br>[redacted] |
| 12:00 – 1:00 PM EST | **Social Hour: Network with Faculty and Staff** |

| | |
|---|---|
| **Thursday, March 3rd** | |
| | **Daily Theme: Share your Hobby** |
| 7:00 – 8:30 AM EST | **Keynote Panel: Emergency and Critical Care** [redacted] |
| 9:00 – 10:30 AM EST | **Keynote: The Role of Biochemical Markers in Monitoring COVID-19 Patients** [redacted] |
| | **Keynote: Molecular Characterization of MDR Bacteria Colonizing COVID-19 Patients in Libya** [redacted] |
| 12:00 – 1:00 PM EST | **Journal Club: Improving Patient Understanding of Treatment Plans** |
| **Friday, March 4th** | |
| | **Daily Theme: Favorite Food** |
| 8:30 – 9:30 AM EST | **Keynote: Telemedicine and home care in the management of severe COVID-19** [redacted] |
| 9:30 – 10:00 AM EST | **Pre-recorded Keynote: Telegeriatrics & the COVID-19 pandemic** [redacted] |
| 11:00 – 11:45 AM EST | **Journal Club: COVID-19 impacts on food supply** |
| 12:00 – 1:00 PM EST | **Social Hour: Pharmacovigilance during the COVID-19 Pandemic** |
| **Monday, March 7th** | |
| | **Daily Theme: School Spirit** |
| 7:00 – 8:30 AM EST | **Keynote Panel: Testing and Diagnostics** [redacted] |
| 9:00 – 10:00 AM EST | **Keynote: A Real Data-Driven Analytical Model for Testing for the Novel Coronavirus Disease** [redacted] |
| 11:00 – 12:00 PM EST | **Journal Club: Optimizing Testing Regimes for the Detection of COVID-19 in Children and Older Adults** |
| 12:00 – 1:00 PM EST | **Poster Viewing** |

#### Tuesday, March 8th

| | |
|---|---|
| | **Daily Theme: Group Participation** |
| 7:00 – 8:30 AM EST | **Keynote Panel: COVID-19 Pathology, Therapy, and Prevention**<br>• Pathophysiology of Platelet Behavior in COVID-19 Infection<br>[redacted]<br>• COVID-19 Therapies<br>[redacted]<br>• Recommended interpersonal distance for Covid-19 prevention<br>[redacted] |
| 9:30 – 11:00 AM EST | **Poster Viewing** |
| 11:00 – 11:45 AM EST | **Social Hour: Working Through Patient Challenges During the COVID-19 Pandemic** |
| 12:00 – 1:30 PM EST | **Workshop: Understanding the Leader in You: Tackling Bias and Imposter Syndrome**<br>[redacted] |

#### Wednesday, March 9th

| | |
|---|---|
| | **Daily Theme: Role Model in Science and Medicine** |
| 7:00 – 8:30 AM EST | **Keynote Panel: Vaccine Considerations for COVID-19**<br>[redacted] |
| 10:00 – 10:45 AM EST | **Poster Viewing** |
| 11:00 – 11:45 AM EST | **Journal Club: Antiviral Drug Discovery: Preparing for the Next Pandemic** |
| 12:00 – 1:30 PM EST | **Workshop: Medical Ethics**<br>[redacted] |

#### Thursday, March 10th

| | |
|---|---|
| | **Daily Theme: Career Goals** |
| 7:00 – 8:30 AM EST | **Keynote Panel: Psychiatric and Mental Health Complications and considerations for COVID-19**<br>[redacted] |
| 9:00 – 10:00 AM EST | **Keynote: Epidemiological Status of COVID-19 in Libya**<br>[redacted] |

| | |
|---|---|
| 11:00 – 12:00 PM EST | **Social Hour: COVID-19 and Mental Health** |
| 12:00 – 1:00 PM EST | **Live Poster Session** |

#### Friday, March 11th

| | |
|---|---|
| | **Daily Theme: Lecture Hall** |
| 7:00 – 8:30 AM EST | **Keynote Panel: The New Normal: Preparing Hospital Operations to Safeguard Lives and Livelihoods**<br>[redacted] |
| 9:00 – 10:00 AM EST | **Keynote: Cardiac Manifestation of COVID-19**<br>[redacted] |
| 11:00 – 12:00 PM EST | **Social Hour: Conference Reflection** |
